# Designing a brief and simple intervention to help young people with type 1 diabetes to live well: protocol for developing a novel intervention with participation from young people

**DOI:** 10.1101/2023.04.20.23288856

**Authors:** Samantha Howland, Jörg Huber, Catherine Aicken

## Abstract

Young people living with type 1 diabetes (T1D) navigate daily complex diabetes related tasks as they take on increasing (and eventually full) responsibility for managing their condition, in addition to developing their lives as independent adults. Alongside the need for careful day-to-day diabetes management, the psychosocial burden and mental health impact and stigma are well recognised. Despite advances in psychological care, many young people with diabetes still experience a greater mental health burden than those without diabetes. This study aims to develop a brief and simple intervention for young people with T1D that will support their wellbeing day to day, as required, and grow their ability to live confidently with their condition that requires lifelong meticulous management. This study will consist of qualitative research and collaboration with young people with T1D and their siblings, friends, and peers to co-create a testable intervention. In Part 1, research interviews will be conducted with young people (16-24 years old) with T1D and, where possible, their siblings/peers to understand the day-to-day challenges of type 1 and what a novel intervention should address. Thematic analysis of interviews will inform the generation of a prototype intervention to take into part two, focus group discussions. Focus groups with (i) young people with T1D and, separately (ii) carers (comprising parents, carers, teachers, specialist nurses). Collaborative principles will be used to review, redesign and evolve the intervention to meet user needs. A blend of narrative and thematic analysis will inform the findings and report.

Insights from Parts 1 and 2 will shape a user-defined and formatively analysed brief and simple intervention and future study design ready for pilot testing. The aim of this part of the research is to maximise acceptability and usability of a testable intervention by the target population. To aim of the future intervention will be to demonstrate effectiveness in helping young adults to live well with T1D.

**METADATA:** This research is funded by a studentship to the first author, provided by University of Brighton as part of its partnership activities with the National Institute for Health Research (NIHR) Applied Research Collaboration Kent, Surrey, Sussex. The funders did not and will not have a role in study design, data collection and analysis, decision to publish, or preparation of the manuscript. The views expressed are those of the authors and not necessarily those of the NHS, the NIHR or the Department of Health and Social Care.

Ethical approval for this study was provided by Cornwall and Plymouth Research Ethics Committee on behalf of the Health Research Authority and Health and Care Research Wales on 22 Aug 2022 (22/SW/0097). The University of Brighton is the Sponsor (protocol number 2022-9871).

## Introduction

It is well recognised that young people transitioning from dependent living as a child or adolescent to independent living as a young adult experience huge changes within many, if not all, aspects of their lives (e.g. peer groups, relationships, geographic location, day-to-day activities at school/ college / university / work, emotional development). Those living with type 1 diabetes need to navigate the added complexity of managing their condition as they take on full responsibility for their diabetes [1]. Life-altering complications are a significant concern with diabetes and studies have shown that risk of developing such complications can be reduced by diligent management and intensive control of blood sugar [2-4]. Regular health review and care is advocated for everyone with diabetes [5-7] to aid prevention, identification and early intervention of any complications of disease.

Accepting the complexities of “adherence to treatment and regular health-related tasks” (willingness, ability, intent, confusion, forgetfulness, definition and measurement of success, etc) more than half of young people with chronic conditions may exhibit so called “non-adherent” behaviour [8, 9] (i.e., intentional or unintentional behaviours related to insulin dosing, glucose monitoring, diet, exercise, or healthcare review that do not align with medical advice). Despite the risks of life-altering complications, diabetes seems to be no exception. Adherence, in the context of young people with diabetes, can be considered as the extent to which a person’s behaviour coincides with medical advice [10] to conduct diabetes-related tasks and includes reacting accordingly to enable or restabilize glucose levels, attend regular appointments with health professionals, etc. Studies have shown that younger adults, including but not limited to transition clinics, are also less likely to attend diabetes health appointments than other age groups [11].

The characteristics of young adulthood are highly variable demographically, reflecting the diverse choices and constraints facing the individual as they explore possible life directions. Young adults typically do not see themselves as adolescents, but many of them also do not see themselves entirely as adults [12]. Their self-perception and self-identity can vary by circumstance and emotion.

For the young adult with diabetes, complex day to day tasks of managing diabetes can therefore be either overlooked in the milieu of other priorities or hidden in order to “fit in” [1]. Where self-management tasks are performed these can cause feelings of stigma, anger, self-resentment, and distress [13, 14] – reasons for which are not always clear.

Several studies have explored ways to enhance self-management of diabetes by young people [15-21] to improve glucose stability and thereby reduce risk of future complications. Interventions (usually in the form of training provided by the patient’s healthcare team[16, 22]) and their evaluation often focuses on clinical outcomes (such as reduction in HbA1C – a biochemical indicator of long term glucose control) as the primary outcome, with psychosocial adaptations reported secondarily [16, 23-25]. These interventions have involved participants ranging between 11 and 24 years. There is evidence, albeit limited, in the medical literature of co-creative/participatory work with young people with T1D in the UK exploring potential adaptations to national health services and support to better serve this patient cohort [26-29].

Recognition of the importance of actively managing psychological and social issues for young people with diabetes is highlighted by the National Institute of Health and Care Excellence (NICE) clinical guideline (NG18) for diabetes (type 1 and type 2) in children and young people [5] which draws attention to the greater risk of emotional and behavioural difficulties in this population. NICE indicates the importance of access to mental health professionals and age-appropriate behavioural interventions or techniques for support.

In preliminary patient and public involvement discussions with young people with diabetes in Summer 2021 they stated that everyone is different and the needs and experiences of living with T1D “vary massively.” They highlighted that they feel that services and support are not really designed for them and do not always focus on their priorities.

The overall aim of the research is to help young people with type 1 diabetes (young people with T1D) to live well with their condition during young adulthood (aged 16-21 years) as they move to independent living, particularly those who are experiencing difficulty or frustration with this lifelong condition. The focus will be holistic rather than targeting HbA1c. Through formative participatory research and utilising design thinking principles, this phase of the project will create a brief and simple intervention with young people and carers that aims to build strength and resilience (optimism, emotional and impulse awareness and control, empathy and connection, self-efficacy and flexible and accurate thinking [30]) so they can manage their diabetes-related tasks and behaviours, allowing them to live well. The intervention will likely be self-guided support and will offer individualised, and readily accessible psychosocial support day to day for young people with T1D. This will be additive to usual care received in structured health and social care settings [5, 31, 32].

This study to design a brief and simple intervention for young people with T1D comprises three parts:

- Part 1: Surveys and semi structured research interviews and design of initial prototype intervention
- Part 2: Surveys and focus group discussions, refining the intervention and planning the pilot study with young people with T1D,
- Part 3: Developing the pilot study of the intervention with insights from Parts 1 and 2.

Secondarily, and after participating in a research activity, each young people with T1D will be asked for feedback about their experience of participating in a research activity about diabetes to gather an indicative assessment on how young people with T1D felt about engaging in research and whether they would recommend others to participate in similar research in the future (based on Net Promotor Score [33]).

The intervention is expected to focus on building up the self-efficacy and self-confidence of young people with T1D through self-guided channels, with an end goal of demonstrating acceptability and usability by young people through formative evaluation by the participants involved in co-creation. The intervention will likely be a toolkit containing self-guided resources in some/all the following areas: handling disclosure of diabetes, asking for diabetes related support, recognizing the impact positive actions, planning for an independent young adulthood, and reflecting on the impact of decisions. Alternatively, or additionally, it could be a series of recommendations on how to design a healthcare service and self-gathered support network to be valuable and helpful to young people with T1D. The content will be such that it can be self-tailored to be meaningful to the individual’s circumstances and self-beliefs. Research interviews and the design of the prototype intervention will be grounded in the behaviour change wheel for designing health interventions [34]. The focus group discussions will apply design thinking principles with the participant groups to tailor the intervention based on feedback. Digital communication will be explored and embedded as appropriate. As reported by Griffiths *et al*, digital communication (text, email, video call) between young people with long-term conditions and their NHS healthcare specialists can improve the health care experience and engagement [35]. The study showed this to be most effective when patients already have a trusting relationship with the healthcare team, and to improve flexible access for support, especially during times of change. This learning will be carried through to the intervention being developed.

The areas of focus identified above have been shaped through a critical literature review covering the previous 10 years, insights from patient and public involvement conversations with young people with T1D and the researcher’s lived experience of T1D.

This protocol describes the planned study and defines the criteria to proceed to pilot testing in a cohort of young people with type 1 diabetes. If effective, the insight-led, novel, brief and simple intervention designed in this work could enhance the lives of young people with diabetes and reduce the mental health burden associated with living with a long-term condition that requires meticulous management.

## Materials and Methods

Through a primarily qualitative design, this participatory research will gather insights and involvement from the target population of young people with T1D. The integration of survey data will add a mixed methods element to inform the design a novel brief and simple supportive intervention will be used to co-develop an intervention with young people with T1D to assist in day to day living well with the condition.

The design comprises three parts as described below and in Figure 1. This adaptive design involves Patient and Public Involvement throughout the running of this study. The formative design is carefully designed to enable active participation with patients and people who know them well in the design of a novel intervention to be pilot tested in the next phase of research.

**Figure 1:**
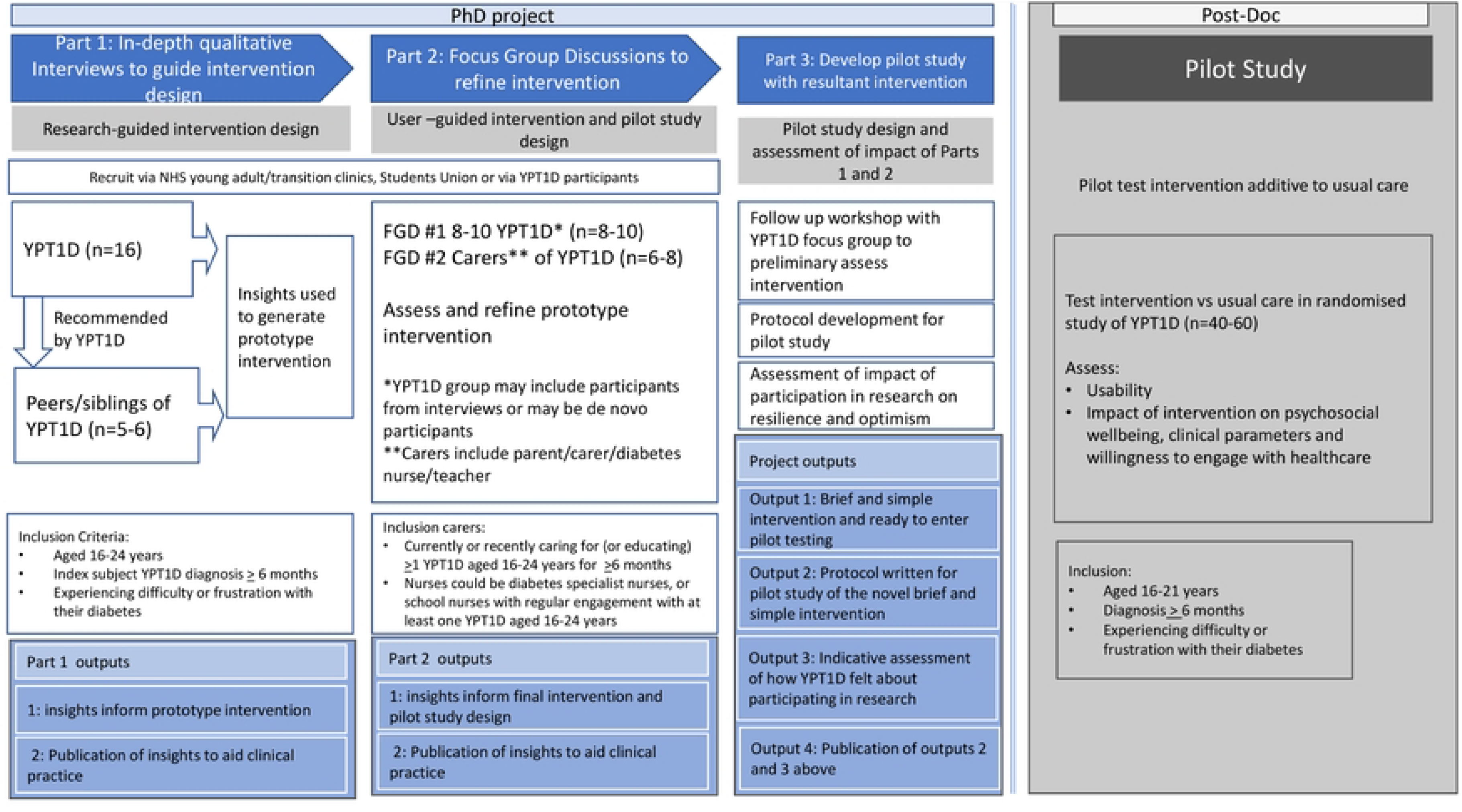
Stages of the Study

### Study Design for Part 1: In Depth interviews and prototype design

#### In-Depth Interviews

Semi-structured interviews will be used to identify the biggest challenges in the participants’ lives at present, what are the most important priorities to young people regarding their diabetes and what they believe would make a difference to help them to live well with their condition as they build their young adult lives.

#### Part 1 (Interviews): Study population

Sixteen patients with type 1 diabetes (age range 16-24 years old), and 5-6 siblings/peers (aged ≥16 years) will be interviewed. This will provide an initial sample size of 21-22 interviews. The sample will be expanded until data saturation is achieved.[36, 37]).

The age range has been chosen to capture input from those near to the target age group for the intervention (16-21 years) to be designed with themes identified in interviews. The lower age limit of 16 years was selected for practical reasons. An upper age limit of 24 years (for young people with T1D) was selected for inclusion in this design phase, to enable reflective thinking from participants on their life experience up to age 21 years.:

i. “Life changes” that impact independence typically occur around the age of 16 years such as school exams, thinking more about university/career/life choices, changing social and education dynamics, gaining independence, learning to drive, etc.
ii. People aged 16+ are legally able to provide informed consent to research participation without parental involvement/consent [38].
iii. Other interventions designed for young people with T1D, such as WICKED (which offered a 5-day self-management education course delivered by health professionals to young people with T1D)[16], similarly define the lower age of the target population as 16 years.
iv. This age group can be reached with targeted recruitment through young adult/transition NHS clinics (which typically manage this age group), which optimises recruitment opportunity through clinic adverts and targeted searching by recruitment sites.

The upper age of 21 years for future testing the intervention was chosen in recognition of the development of independence of young adults in their early twenties and the topics may be perceived as less relevant for older individuals.

Siblings/peers are included to provide a close external perspective of the issues faced by young people with T1D, additive to self-reflections. An upper age limit was not set for siblings/peers for inclusivity of a variety of external opinions and in recognition that siblings’ and peers’ ages can be close or far apart.

#### Part 1 (Interviews): Inclusion/exclusion criteria

Inclusion and exclusion criteria are shown in Figure 2.

**Figure 2:**
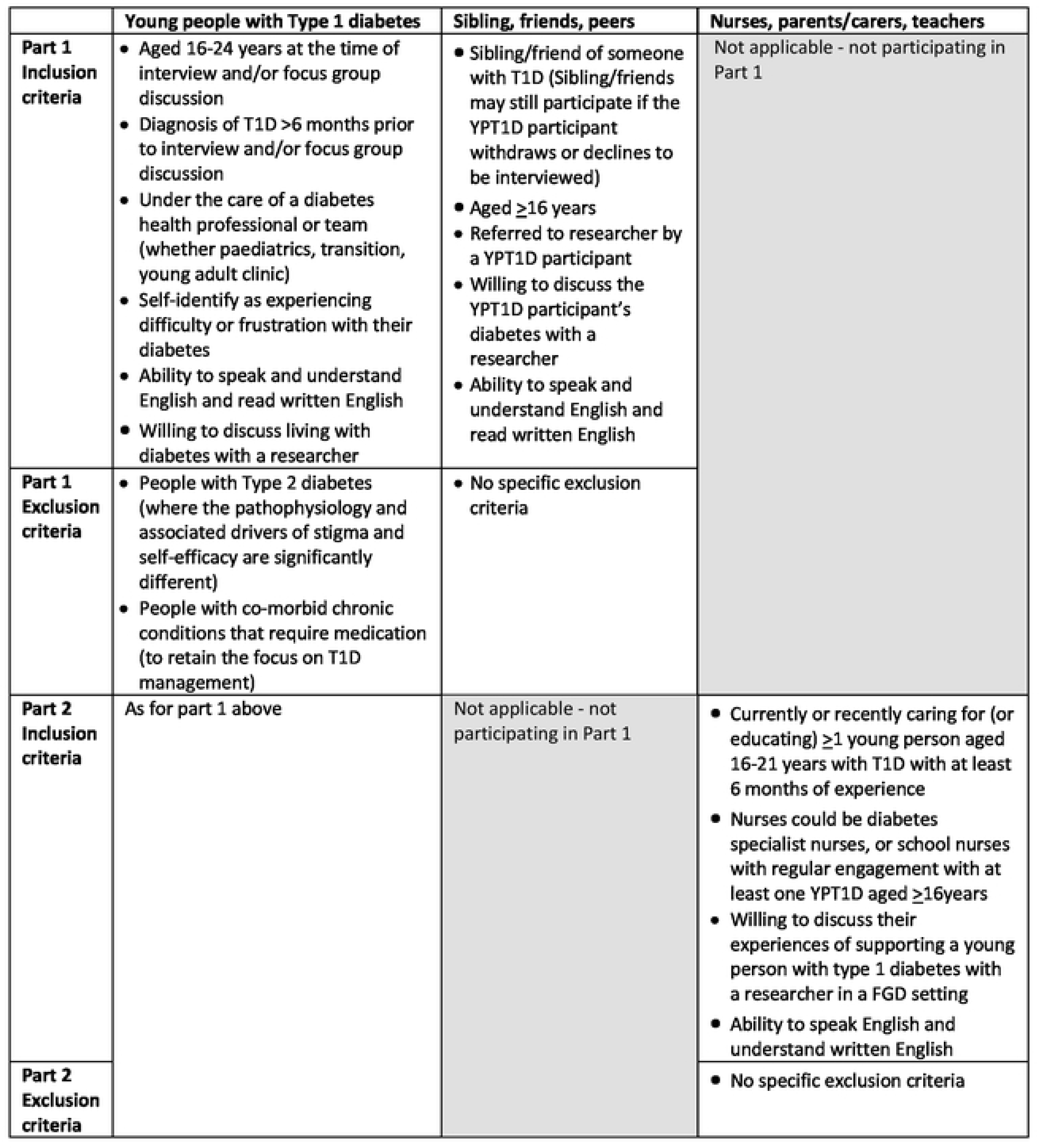
Inclusion and Exclusion Criteria

#### Part 1 (Interviews): Recruitment

young people with T1D participants will be recruited from two (or three) National Health Service (NHS) clinics in England for young people with T1D and through adverts placed on the University of Brighton’s Student Union website. A participant information sheet (PIS) will be provided by the researcher to all potential participants. young people with T1D will be provided with a PIS for their own involvement plus a separate PIS to pass on to their siblings/peers. Clinical nurses may also provide the PISs to their patients in clinic. The young people with T1D will have the authority to choose whether to pass on to their sibling/peer or not. The PIS for siblings/peers will also be provided to the sibling/peer by the researcher when contact is established. PISs will be available in printed form, via email and via the closed group Instagram study page.

Peers/siblings will be identified by asking the young people with T1D participants with T1D during recruitment whether they have a sibling or friend meeting the inclusion criteria who may also be willing to participate in a research interview for this study. There will be no obligation to identify a peer/sibling to take part in the study. It will be explained to each individual participant that they can make their own independent decision about whether to take part. This recruitment route was chosen to provide power to the primary participants (the young people with T1D) to enable them to control whether or not a sibling or peer will participate in the study.

The researcher will ensure each participant can ask questions about the study and will go through the informed consent process and use a separate informed consent form for each individual participant prior to any research activity with that individual. Participants will be free to withdraw consent at any point at which point no further involvement in the research activity would be asked of them.

An informed consent form will be given to each participant to read, review and sign (if research takes place in-person). For remote research activities, informed consent may be captured verbally in a non-audio recorded conversation and documented by the researcher.

Each participant will be at least 16 years old and able to self-consent to participate in research in accordance with NHS Health Research Authority guidance "Research Involving Children [updated Sept 2021]". Parental consent will not be required.

All participants will be offered compensation in recognition of the time given to the interview. This includes young people with T1D, and siblings/peers. Any support person/buddy (if requested by participants) will not receive compensation.

#### Part 1 (Interviews): Sampling

After capturing informed consent, young people with T1D participants will be purposively sampled from across the age range and will include a mix of genders. Self-selected volunteers and potential participants identified by site nurse(s) will be purposively sampled using a combination strategy after completion of a Demographics Questionnaire. First stratified for gender mix (with a view to achieve parity of male: female participants, but also inclusive of other genders e.g., non-binary) and then stratified for variation in age with a view to include opinions from YPTID aged 16-19 years and 20-24 years in roughly equally numbers. Chronological age will be used as a proxy for maturity and independence, and in anticipation that patients’ needs from health services and family/friends may be different based on age. Anticipated numbers of young people with T1D participants are shown in Table 1 Anticipated sampling quotas for young people with T1D participants. By the nature of the voluntary recruitment strategy, it is accepted there is an element of opportunism in the sampling and it will be accepted that the final group may not be equally distributed. Siblings/friends/peers will not be stratified for inclusion, but basic demographics will be collected to understand age and connection to young people with T1D.

**Table 1.** Anticipated sampling quotas for young people with T1D participants

|  | Female young people with T1D | Male young people with T1D | total |
| --- | --- | --- | --- |
| 16-19 | 4+ | 4+ | 8+ |
| 20-24 | 4+ | 4+ | 8+ |
| total | 8+ | 8+ | 16+ |

#### Part 1 (Interviews): Data Collection

Participants with T1D will be invited to complete the World Health Organisation – 5-item wellbeing index (WHO-5) [39] and Problem Areas in Diabetes (PAID) [40] questionnaires via a direct link onto a secure questionnaire per participant on JISC Online Surveys prior to conducting qualitative semi-structured interviews on a one-to-one basis. Interviews with nominated peer/siblings will be conducted separately. Joint (dyadic) interviews with the young people with T1D and their sibling/peer will be accepted if requested by the young people with T1D.

A support person/buddy will be permitted if requested by the young person. This will be made clear on the PIS. The buddy will be provided with an information sheet to explain their role - to ensure that the interview is conducted sensitively and respectfully and to support any needs they may have. They will not be considered as part of the research interview. Buddies’ views will not be sought nor reported by the interviewer during the interview, and through the information sheet they will be dissuaded from commenting on or influencing the young people with T1D.

Interview guides will provide a framework for individual qualitative interviews that have been written with a blend of questions around specific areas of living with diabetes – such as at school/work/university and in interactions with family, friends, and significant others – as well as broader questions around day-to-day life. The intent of this approach is to encourage the participant to open-up through storytelling and thus for the researcher gain a deep understanding of the lived experience and challenges in different parts of life in their own words. The interviewer will seek to understand what they would find helpful (in a non-clinic setting) to address some of the challenges they face with diabetes. Interview guides will evolve iteratively per standard qualitative research practice. (Interview guides are provided in Supplementary Information).

Visual aids will be available (see Supplementary Information) during the interviews to help participants to articulate emotions and feelings associated with their experiences. These will be used at the discretion of the researcher where a participant struggles to articulate their thoughts/feelings or is not forthcoming with feelings or emotional impact. The stimuli were developed from words used by PPI participants, and distilled from literature relating to basic emotions [41] plus the researcher’s own experience. Basic internet searching for copyright free images related to specific emotions was used to create the illustrations.

Confidential individual or dyadic interviews will be held online with a secure VoIP (Voice over Internet Protocol) platform (such as Teams (preferred), Skype, or FaceTime) depending on the participant’s preferred channel for a real-time video call. If VoIP technology cannot be used (eg due to data charge concerns from participants or no webcam enabled device), interviews will take place by telephone. This remote approach is based on the anticipated continued caution in face-to-face interactions owing to the COVID-19 pandemic and aims to enable participation of a wider demographic of participants who previously may not have been willing or able to attend an in-person interview. The researcher will follow best practice for conducting qualitative research interviews online/by telephone effectively. This includes building rapport prior to “meeting” through email/messages, assessing non-verbal cues, and understanding the lived experience in a home/natural environment. It also includes logistical elements such as using private physical and online spaces for the conversation to protect participants’ privacy as well as letting them know that they can pause the interview if they are interrupted (since the researcher may not be able to see if another person has entered the room) [42].

After their interview, young people with T1D participants will be asked how they felt about participating in the research and whether they would do something similar or recommend a friend to participate in similar research in the future. A “Thank You” communication will be sent to all participants along with the compensation after individual interviews.

#### Part 1 (Interviews): Data Management and Analysis

Interviews will be conducted, recorded, and transcribed verbatim, manually by the researcher. A password protected digital voice/video recorder will be used along with Teams automatic transcription option, assuming consent is granted for audio recording and that Teams is used as the platform for the conversation. Transcripts will be anonymised during data familiarisation and coded thematically [43] using NVivo 12 (QSR International, March 2020). Acceptance of recording the interview will be part of the informed consent procedure for participants to ensure that detailed and thorough interpretation can be made of the discussions. Any citation used from the interview will be anonymised, so participants’ privacy is protected. Transcripts will be analysed with thematically coding. Four interviews will be selected at random, transcripts checked, and coding reviewed by a second member of the research team to assess alignment in interpretation (representing ∼20% of the sample size). Any discrepancies that cannot be resolved by the two researchers will be moderated by a third member of the research team. If this situation arises, additional coded interview(s) will be reviewed to ensure alignment on interpretation. The number of reviews of coded interviews will continue until consensus is reached regarding interpretative themes.

Themes/codes will be focused on areas of concern that impact wellbeing and self-belief for young people with T1D to understand context and social influence of the areas of concern. Themes from the coded data will be assessed for amenability to change and used to identify recommendations that will shape an initial idea for a brief and simple intervention for young people which will be used as an initial concept in Part 2 where focus groups will review, rebuild and refine the intervention using a blend of co-creation principles [34, 44, 45].

Findings from WHO-5 and PAID questionnaires will be reported. Indirect comparison of findings with other similar cohorts (young adults with type 1 diabetes for ≥6 months) will be conducted to show, indicatively, whether the sample is representative of other populations with type 1 diabetes matched as closely as possible for age and duration of diabetes). PAID responses will be used to identify other problematic areas for participants associated with diabetes that may not have surfaced in the interviews.

#### Part 1 (Interviews): Outputs

From the work in part 1, skills or areas (themes) that young people identify as high priority to change that would improve to give them confidence to live well will be identified. These high priority themes will be assessed for ability to change at the individual level by taking learnings from other programmes such as TEENCOPE (USA) [15], WICKED (UK) [16] and BITES (UK, brief intervention although not specific to young people)[25]. High priority themes amenable to change that may reduce the barriers to diabetes adherence through building self-efficacy and coping mechanisms will be built into a brief and simple prototype intervention for young people with T1D.

Following the Behaviour Change Wheel approach to designing interventions [34], potential actions/changes that could improve the targeted behaviour of self-efficacy and coping strategies will be generated by the research team and assessed for (a) impact, and (b) likely of behaviour change, (c) effect of change on other behaviours and (d) ability to measure change using a 0-4 ranking scale for each element. The highest-ranking potential actions will become the focus of a prototype intervention.

The prototype intervention, in the form of a PowerPoint representation or infographics, will be created by the researcher using insights gained in the Research Interviews will be taken into Part 2 - focus group discussions.

#### Study Design for Part 2: Focus group discussions (FGDs)

Ideas and opinions from the participants of the focus groups will be utilised to refine a prototype toolkit for young people with type 1 diabetes as designed at the end of Part One.

#### Part 2 (Focus Group Discussions) - Population

Two focus groups will be convened to review, rebuild and refine the intervention concept (prototype) created by the researcher using themes identified in Part 1.

i. One focus group will consist of young people with T1D– the intended users of the intervention - ideally in a dyad with a self-nominated peer (n=8-10)

- Participants will invited following part 1 research interviews (if they have agreed to participate in further research). It will be permitted to extend the invitation to FGD to other young people with T1D who were unconnected to the research interviews in Part 1 (e.g. to increase the numbers of participants if Part 1 interviewees decline further involvement).
ii. The other focus group will consist of diabetes specialist nurse(s), teacher(s) and parents/carers. (n=6-8)

- Participants may or may not be connected to the young people with T1D in FGD (i)

#### Part 2 (Focus Group Discussions): Inclusion/exclusion criteria

Inclusion and exclusion criteria are shown in Figure 2.

#### Part 2 (Focus Group Discussions): Recruitment and sampling

Recruitment tactics described above for Part 1 will be mirrored for the FGD with young people with T1D. Participants from Part 1 will be allowed to participate in Part 2 but additional recruitment of new young people with T1D may be considered if required for numbers.

Parents/carers/teachers will be sourced by asking the young people with T1D participants with T1D during Part 1 research interviews whether they would be willing to recommend one of their parents/carers and/or a teacher who may also be willing to participate in a focus group discussion for this study.

Anticipated numbers of parents/carers, teachers and diabetes nurses are given in Table 2.

**Table 2.** Anticipated quotas for Focus Group Discussion with carers

| Focus Group Discussion participants | Target number of participants |
| --- | --- |
| Parent or carers | 2+ |
| Teacher | 2+ |
| Diabetes nurse | 2+ |
| **Total** | 6-8 |

All activities will be voluntary, and individuals will provide consent for their own participation for each applicable part of the research and following the principles described above in Part 1. There will be no obligation for young people with T1D to identify a parent/carer/teacher to take part in the study. It will be explained to each individual participant that they can make their own independent decision about whether to take part. The researcher will ensure each participant can ask questions about the study and will go through the informed consent process and use a separate informed consent form for each individual participant.

Recruitment for the focus groups will include asking Part 1 interview participants with young people with T1D whether they would like to join a FGD. The recruitment period will extend to 6 months post interviews if additional participants need to be found. FGDs will occur after the completion and analysis of Part 1.

Participants will be offered compensation in recognition of the time given to the focus group discussion. If the meeting is held in-person, reasonable travel expenses will be recompensed (eg standard class rail fare, parking, mileage at prevailing *gov.uk* rate for business travel).

The number of participants has been selected to enable the voice of each participant to be heard in the FGDs whilst still enabling a range of opinions to be captured. Focus group (ii) comprises fewer individuals as the intent is to gather more general reflections from the perspective of parents/carers/teachers/nurses as opposed to specific feedback on acceptability and content from the anticipated users of the prototype intervention from the young people with T1D FGD (i).

As for Part 1, a participant information sheet will be provided to each potential participant with the opportunity to ask questions. Informed consent will be obtained by the researcher and will be recorded from all participants by the researcher. Consent can be freely withdrawn at any point. Due to the nature of group discussions and difficulties identifying individual speakers, it will not be possible withdraw individual data gathered during FGD up to the point of consent withdrawal (however they could leave the discussion part-way through).

#### Part 2 (Focus Group Discussions): Data Collection

Prior to attending the FGD (FGD i), new participants with T1D will be asked to complete a demographics questionnaire, the WHO-5 [39] and PAID [40] questionnaires using a secure link to JISC Online Surveys. The questionnaires are provided in **Error! Reference source not found.** Face-to-face groups will be planned but with a back-up option of virtual meetings (refer to Part 1 for VoIP platform information). Delivery will be governed by any COVID-19 (or similar) restrictions on meetings and associated public health, NHS and University guidelines.

Insights will not be shared between focus groups. Both will be treated as *de novo,* recognising that perspectives on how to support transition to adulthood could be very different for young people compared to carers.

Carers’ perspectives (FGD ii) will be explored to capture close external perspectives and ideas.

An online web forum may be used (e.g., for co-creating material in a focus group discussion). Access will be restricted to participants and supervisors only. Informed consent will be obtained from all participants to work on this shared platform prior to use. Participants may choose to use pseudonyms if preferred.

FGDs will be recorded and transcribed verbatim apart from anonymization as described in Part 1 (for research interviews).

At the start of the FGD, Ways of Working in the Workshop will be discussed and agreed by all participants. This will include as a minimum (a) respect for the privacy of workshop discussions, (b) respect of others’ opinions and ideas, (c) allowing everyone to speak and contribute their individual ideas. The outlined principles (see figure 2) may be tailored by the participants.

Using design thinking principles [45], focus groups will be convened with young people with T1D and, separately carers of young people with T1D, to review the prototype, comment and provide ideas for improvement or change.

Design thinking is a qualitative methodology that is centred on input from target users, more commonly used in business and technology than healthcare for ill-defined complex problems. The approach uses insights from potential users and environmental sources to create new ideas and solutions. The design thinking model comprises five phases – empathy for the issue, definition of the problem, ideation around potential solutions, creation of a prototype to address the problem, and testing [46]. Iteration is used for refinement as required. This participatory method was chosen to ensure that the voice and desires of the end users are fundamental to the design of the intervention. It is a less structured methodology than the Behaviour Change Wheel (BCW) [34] however elements of BCW will also be utilized as defined below.

Using the BCW for defining the problem, participants will be asked to reflect on the themes identified in the research interviews to define the problem(s) identified by interviewees in behavioural terms (what behaviour are we seeking to change, where does it occur, who is involved). A COM-B (’capability’, ’opportunity’, ’motivation’ and ’behaviour’) approach will be taken to structure description of what needs to happen for the target behaviour to change and to articulate whether there is a need for change. To ideate potential solutions, the prototype intervention will be shared with the participants who will be asked to review whether it meets the areas of focus outlined above. Questions will be addressed and areas to change/new ideas will be captured.

Participants will also be invited to feedback and comment on an initial proposal of a pilot study (including target population for the study and potential assessment questionnaires from an *a priori* list). The intent is to understand what would likely be acceptable, usable, and valued by the participants in the future pilot study. Suggestions for other assessments by the participants will also be considered during the focus group discussion as long as the areas of interest can be captured using validated questionnaires. The intent is to design the intervention and future pilot study so that it will likely be acceptable and usable by the participants. Suggestions for other outcomes in a pilot study that would be important to young people will also be considered during the FGD discussion.

At the end of the focus group discussion with young people with T1D the participants will be asked if they would be willing to look at the resultant intervention incorporating their ideas as formative testing prior to taking forward to pilot testing. If they consent, the participants will be asked to consent to further participate via email, message, video call, telephone call or face to face (the meeting set up would be their choice) where they would be able to review and ask questions and offer suggestions for further modifications to the intervention on a one-to-one or small group basis. Agreeing to further engagement is not a pre-requisite of attending the focus group discussion. A separate consent form will be used for FGD follow up.

After their participation, as in Part 1, young people with T1D participants will be asked how they felt about participating in the research and whether they would do something similar or recommend a friend to participate in similar research in the future. A “Thank You” communication will be sent to all participants along with the compensation after the focus group discussion.

#### Part 2 (Focus Group Discussions): Data Management and Analysis

If conducted in person, focus group discussions will be recorded using a password protected digital voice recorder. If the meeting(s) occurs onscreen, the Teams automatic transcription option will be used in addition to the voice record, assuming consent is granted for audio recording.

A narrative of learnings will be generated to summarise the FGDs and recommendations within [47].

Findings from WHO-5 and PAID questionnaires will be reported as in Part 1.

#### Part 2 (Focus Group Discussions): Outputs

Insights and recommendations from FGDs will be taken forward by the researcher into Part 3 where the intervention will be rebuilt or refined ready to pilot test in a randomised study. A pilot study protocol will be created.

#### Study Design for Part 3: Building the intervention ready to pilot test and defining the pilot study protocol

The initial prototype designed with insights from the research interviews in Part 1 will be reviewed and adapted by the researcher based on the insights from the FGDs in Part 2.

After adaptation of the intervention, the researcher may return to the FGD participants (pending their consent) to show them the resultant intervention prior to taking forward to pilot testing. The participants will be able to review and ask questions and offer suggestions for further modifications. Formative evaluation will be conducted with the participants through fielding (a) the user experience questionnaire [48], and (b) a bespoke multiple choice questionnaire about the intervention to assess acceptability and usability of the of the intervention. Participants will be invited to complete questionnaires via a direct link onto a secure questionnaire via JISC Online Surveys per participant. Using insights from the FGDs, a protocol for testing the final intervention in a randomised pilot study will be created by the researcher.

#### Data handling and Data Protection

Participants will be assigned a study number by the researcher which will be used in all documentation relating to the research (eg interview transcripts, FGD participant lists, questionnaire documents, etc). General Data Protection Regulations (GDPR) will be adhered to ensure data privacy and appropriate data handling.

### Safety Considerations During Data Collection Activities in Parts 1 and 2

In depth interviews and group discussions about living with diabetes that may cover topics that young people and/or parents may be sensitive or frustrated about. This may generate emotional response and/or distress. An Issues Management guide will be included in each interview/focus group discussion activity guide so that such situations can be handled appropriately at the time to ensure the wellbeing of the participant.

- For example, an interview may be paused, stopped, or rescheduled if needed.

Disclosure of anything life threatening or that may impact their immediate safety, physical or mental health of the participants or others would be handled urgently and referred for appropriate support within 24 hours.

Living with diabetes has an omnipresent risk of occurrence of out-of-range blood glucose - either hyper- or hypoglycaemia. This may present as acute unwellness and would likely require urgent attention by the participant to correct blood glucose.

- During Part 1 (research interviews) if any participant became unwell during the interview (eg due to hypoglycaemia) the interview would be stopped. The participant would be asked if they would like to re-engage in the interview once they had fully recovered.
- During Part 2 (focus group discussions) if any participant became unwell during the discussion (eg due to hypoglycaemia) they would be able to leave the discussion. The discussion would be paused to ensure the wellbeing of the individual but may carry on either with or without the affected individual. Treatment for hypoglycaemia (glucose gel, glucotabs, carbohydrate-based snacks) will be available in the room if required.

People living with diabetes may be at higher risk of transmissible infection, or sequalae of infection, than other people. Considering COVID-19 or similar pandemic/endemic public health infection risk:

- Part 1 research interviews will be conducted via video call or telephone call to avoid risk of transfer of infection and to enable broader participation.
- Part 2 focus group discussions are intended to be face-to-face and will be conducted with COVID-19 safe procedures in place (ventilation, hand sanitiser provided, face masks provided if required by Public Health guidance).

`o` If recommended by national, local or organizational guidelines, FGDs will be able to be conducted online via video conference to avoid risk of infection.
`o` The more conservative approach will be taken if guidelines differ.

### Patient and Public Involvement (PPI)

The adaptive design of this study involves PPI throughout the running of this study and is described throughout this article.

## Discussion

### Main contributions of the study

The learnings from this project will be used to design a user-informed, testable brief and simple intervention that will be taken forward into pilot testing where the intervention will be assessed in a randomised sample of young people with diabetes (aged 16-21 years) in England.

Input from the FGD groups will inform the pilot study design and how to assess success and impact on wellbeing, clinical management of diabetes and willingness to engage with healthcare. A prototype design of a pilot study and the proposed outcomes will be shared with the group and the group will be asked to reflect on, and make recommendations whether, the questionnaire-based outcomes are appropriate, and whether they would like to consider other outcomes within the study. This input will provide preliminary evidence of the acceptability and feasibility of the trial design.

### Strengths and Limitations

The participatory approach with young people with T1D aims to enable identification of key areas for the intervention to focus on. Their input will inform the style and narrative of the intervention to optimise opportunity for success. Involving young people in interviews and FGDs may also have some psychological benefit, with participants possibly benefitting from the primary focus on their own opinions, sharing experiences and feeling empowered through being treated as valued consultants.

Giving the young people with T1D control about who, if anyone, they nominate to take part and whether they are interviewed together/individuality in Part 1 is similarly intended to give shared control and primacy to their opinions. In doing so, it is anticipated that rapport and respect will be more readily built, and the participants will engage deeply in the research providing insights and creative ideas. This level of input and review in FGDs aims to optimise the usability and acceptability of the intervention to the target population.

Within the qualitative formative research there is some potential to explore the influence of past specific experiences (recent or otherwise) that have led participants to the self-beliefs and priorities that they discuss in the interview. However, this is limited compared to, for example, a longitudinal qualitative study on the evolution of diabetes-related beliefs. These are beyond the scope of this research. Evolution of diabetes-related beliefs through childhood and adolescence will be explored through review of other studies.

Recall bias may present where participants may focus on one particularly challenging incident that has shaped their thoughts on that day. Recency and saliency (and perceived social desirability/acceptability) are expected to influence topics brought up by the participants. Encouragement of reflective thinking and introduction of time-bound questions (eg can you remember how you felt 6 months before xxx happened. What was it like then?), along with structured questions around different life areas will be used to aid recall and widen the focus beyond a few key incidents or memories. To encourage discussion of topics that challenge social desirability/acceptability, the researcher will focus on building trust and rapport early in the engagement. Confidentiality will be assured (as per informed consent).

The potential for selection bias of young people participating in any part of this research is an inherent bias/limitation to any study based on voluntary participation. Those young people who chose to participate may differ in an important way from those who chose not to participate.

By the conversational nature of Part 1 (research interviews) and Part 2 (FGDs), this research may better capture the views, preferences, and experiences of participants who are willing and able to speak out and share ideas (anticipated to be more extrovert/confident/articulate) than quieter, solitary or shy individuals. Engaging with teachers/carers/nurses in Part 2 (FGDs) will provide an indirect opinion about likely acceptability of the prototype intervention by more introvert/shy/quiet/less articulate young people with T1D.

A further selection bias could be driven by the sites (NHS and university) that participate in, and support recruitment of, young people into the study. Not only from a socio-economic demographic perspective but also that the beliefs and priorities of the young people included may well have been shaped by the skills and interactions with their healthcare providers, their friends, their school environment, etc. Demographics and services are known to be different around the country so it is reasonable to assume that patients’ self-beliefs and priorities may also differ in different geographies.

Owing to the conversational nature of the formative research, the inclusion criteria for this study is reliant on participation of those who can speak and understand English. This potentially excludes other people who might also benefit from this intervention. Whilst a limitation at this formative stage, translation and non-written adaptations may be considered as a development in future research.

The other limitations identified in this section will be explored in further research that would widen the reach and usability pending successful completion of the pilot study.

### Future Direction

Insights and learnings from this work will be shared with participants and submitted for publication to raise awareness of the issues and priorities of young people with diabetes.

The design of the final intervention (output from Part 3) will be taken forward into pilot testing (pending future ethical approval) and will be published in association with the pilot study. If the pilot study shows preliminary evidence of effectiveness of the intervention, it will be scaled to a larger population in a randomised controlled trial and health economic evaluation to assess (i) the scale and durability of improvement that is achievable in different age groups, and (ii) the health economic impact of the intervention.

Depending on the transferability of the priority areas identified in this study, extension of the intervention to other chronic conditions of adolescence/young adulthood which require some element of self-management will be explored (eg maturity-onset diabetes of the young (MODY), cystic fibrosis, juvenile idiopathic arthritis, haemophilia).

If effective in testing, this intervention and the insights from this work could enhance the lives of young people with diabetes and reduce the mental health burden associated with living with a long-term condition that requires meticulous management.

## Data Availability

Deidentified research data will be made publicly available when the study is completed and published.

## ACKNOWLEDGEMENTS

Thanks are due to academic colleagues, clinicians and patients who have contributed to the design of this programme through provision of feedback on the protocol and patient materials.

